# Evaluation of transplacental transfer of mRNA vaccine products and functional antibodies during pregnancy and early infancy

**DOI:** 10.1101/2021.12.09.21267423

**Authors:** Mary Prahl, Yarden Golan, Arianna G. Cassidy, Yusuke Matsui, Lin Li, Bonny Alvarenga, Hao Chen, Unurzul Jigmeddagva, Christine Y. Lin, Veronica J. Gonzalez, Megan A. Chidboy, Lakshmi Warrier, Sirirak Buarpung, Amy P. Murtha, Valerie J. Flaherman, Warner C. Greene, Alan H.B. Wu, Kara L. Lynch, Jayant Rajan, Stephanie L. Gaw

## Abstract

Studies are needed to evaluate the safety and effectiveness of mRNA SARS-CoV-2 vaccination during pregnancy, and the levels of protection provided to their newborns through placental transfer of antibodies. We evaluated the transplacental transfer of mRNA vaccine products and functional anti-SARS-CoV-2 antibodies during pregnancy and early infancy in a cohort of 20 individuals vaccinated during pregnancy. We found no evidence of mRNA vaccine products in maternal blood, placenta tissue, or cord blood at delivery. However, we found time-dependent efficient transfer of IgG and neutralizing antibodies to the neonate that persisted during early infancy. Additionally, using phage immunoprecipitation sequencing, we found a vaccine-specific signature of SARS-CoV-2 Spike protein epitope binding that is transplacentally transferred during pregnancy. In conclusion, products of mRNA vaccines are not transferred to the fetus during pregnancy, however timing of vaccination during pregnancy is critical to ensure transplacental transfer of protective antibodies during early infancy.

## Introduction

Growing evidence has shown that pregnant individuals are at higher risk for SARS-CoV-2-related morbidity and mortality^1-4^. Despite this, vaccination uptake by pregnant individuals has been slower than the general population^5^, in part because of maternal concern of adverse effects on the embryo or fetus, even with strong consensus recommendations for COVID-19 vaccination prior to or during pregnancy from several medical societies^6^. Pregnant individuals were excluded from initial vaccine trials, and complete data on safety, efficacy, optimal timing of the vaccine in pregnancy, or its impact on the fetus has been delayed^7^, which may impact individual medical decision making. Current COVID-19 vaccines fully approved and under emergency use in the United States include the mRNA vaccines BNT-162b2 and mRNA-1273, which target the SARS-CoV-2 Spike protein and stimulate protective immune responses ^8,9^. In addition to protecting the mother against severe disease, vaccination during pregnancy may protect the newborn through passive transfer of maternal immunoglobulin. SARS-CoV-2 infection and vaccination during pregnancy produces an IgG response that is transferred to the fetus^10-16^. Evidence of newborn protection might help address maternal concerns about adverse effects. However, detailed studies of the transplacental transfer of vaccine products and vaccine-related antibody dynamics, functional properties, and persistence during infancy of transferred SARS-CoV-2 antibodies are needed to provide such evidence.

We examined the transplacental transfer of mRNA vaccine products and humoral responses using samples from pregnant individuals and their infants vaccinated with either BNT-162b2 or mRNA-1273 mRNA vaccine during pregnancy.

## Results

### Cohort

We evaluated 20 pregnant individuals who received COVID-19 mRNA vaccines during pregnancy and their infants. Participants were vaccinated between December 2020 and April 2021. Gestational age at first dose ranged from 13 weeks to 40 weeks (mean 31.2, SD 5.9 weeks). Nineteen participants delivered live, singleton infants between January 2021 through April 2021 at gestational ages ranging from 37.4 to 41.1 weeks (mean 39.2, SD 1.1 weeks). One participant who was vaccinated at 13 weeks had a termination of pregnancy due to a lethal skeletal dysplasia of genetic etiology at 20.4 weeks. Eight participants received BNT-162b2 (Pfizer-BioNTech) and twelve received mRNA-1273 (Moderna) vaccines. Eighteen participants received both vaccine doses prior to delivery, and two participants received the second dose after delivery. The time from first mRNA vaccine dose ranged from 6-97 (mean 51, SD 24.3) days prior to delivery, time from the second dose ranged from 2-75 (mean 32, SD 21.3) days prior to delivery, and in two participants 15 and 21 days after delivery. No participants received a 3^rd^ dose prior to delivery. Infants born to vaccinated mothers were followed up at convenience time points ranging from age 3 weeks to 15 weeks of life (mean 8.3, SD 3.2). Further demographic data is detailed in Table S1.

### Vaccine mRNA products do not cross the placenta

To determine the transplacental transfer of mRNA vaccine derived products, we examined available maternal blood at delivery, placenta tissue, and cord blood for Spike protein by Western blot and vaccine mRNA by PCR. All available delivery samples (maternal blood, placental tissue, and cord blood) were negative for Spike protein by Western blot (Supp Figure 1, Supp Table 3) and did not have detectable levels of vaccine mRNA by PCR (Suppl Table 3). Together, this indicates that products of mRNA vaccination do not reach the fetus after vaccination during pregnancy at readily detectable levels.

**Figure 1.**
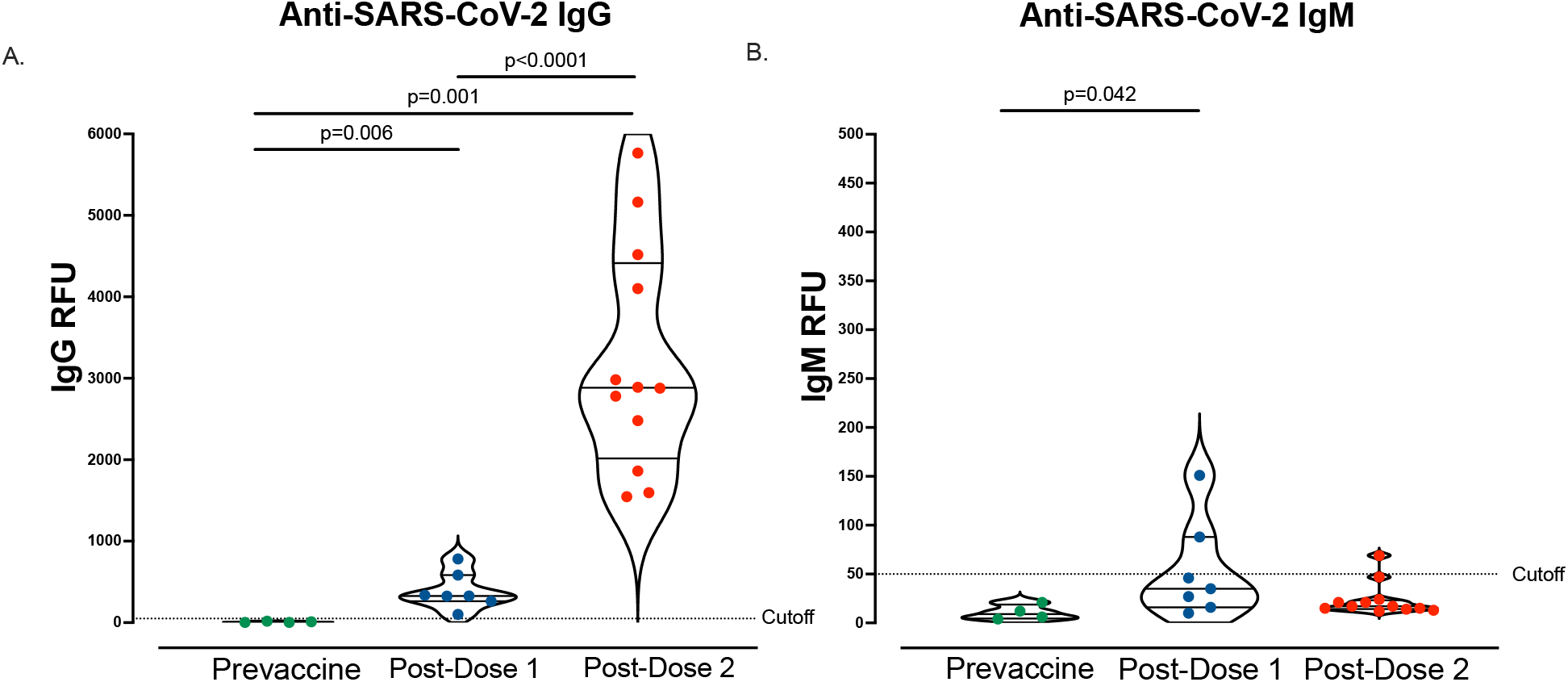
Anti-SARS-CoV-2 IgG and IgM antibody responses following vaccination. A. Maternal plasma anti-SARS-CoV-2 IgG antibody relative fluorescence units (RFU) levels prior to vaccination (n=4), 3-4 weeks post-dose 1 (n=7), and 4-8 weeks post-dose 2 (n=12). B. Maternal plasma anti-SARS-CoV-2 IgM (RFU) levels prior to vaccination (n=4), 3-4 weeks post-dose 1 (n=7), and 4-8 weeks post-dose 2 (n=12). Wilcoxon rank-sum testing. Data represent median ± quartiles, two-sided *p* values were calculated for all test statistics.

### mRNA vaccination in pregnancy leads to a robust antibody response

Similar to prior studies^14,15,17^, we found that mRNA vaccination during pregnancy led to an increase in anti-SARS-Cov-2 IgG following dose 1 (n=7, mean 388.6, SD 224.8 RFU) and an even further robust increase after vaccination dose 2 (n=12, mean 3214, SD 1383 RFU). Anti-SARS-CoV-2 IgM (n=7, mean 53.3, SD 50.2 RFU) was detected in two maternal participants following dose 1, but only 1 participant following dose 2 (n=12, mean 23.8, SD 17 RFU, Fig 1).

### Vaccine induced anti-SARS-CoV-2 IgG and neutralizing antibodies are transplacentally transferred

We then evaluated the transplacental transfer of maternally derived anti-SARS-CoV-2 IgG antibodies to their infants. Maternal blood at delivery was available in 19/20 participants and cord blood was available in 17/20 participants. Anti-SARS-CoV-2 IgG was detectable in 94.7% (18/19) of maternal blood samples at delivery (mean 3235, range [10, 7811] RFU). Anti-SARS-CoV-2 IgG was detectable in 88.2% (15/17) cord blood samples (mean 2243, range [2, 4959] RFU). One participant received one mRNA vaccine dose 9 days prior to delivery, and both the maternal and cord blood were negative for IgG at the time of delivery. Another participant received two doses of mRNA vaccine (23 and 2 days) prior to delivery and maternal blood was positive at 55 RFU (positive cutoff >50 RFU), however cord blood IgG was negative (Figure 2A). Maternal and cord blood anti-SARS-CoV-2 IgG levels were moderately correlated, but not statistically significant (p=0.074, R_s_=0.446, Fig 2A). All cord blood samples were anti-SARS-CoV-2 IgM negative.

**Figure 2.**
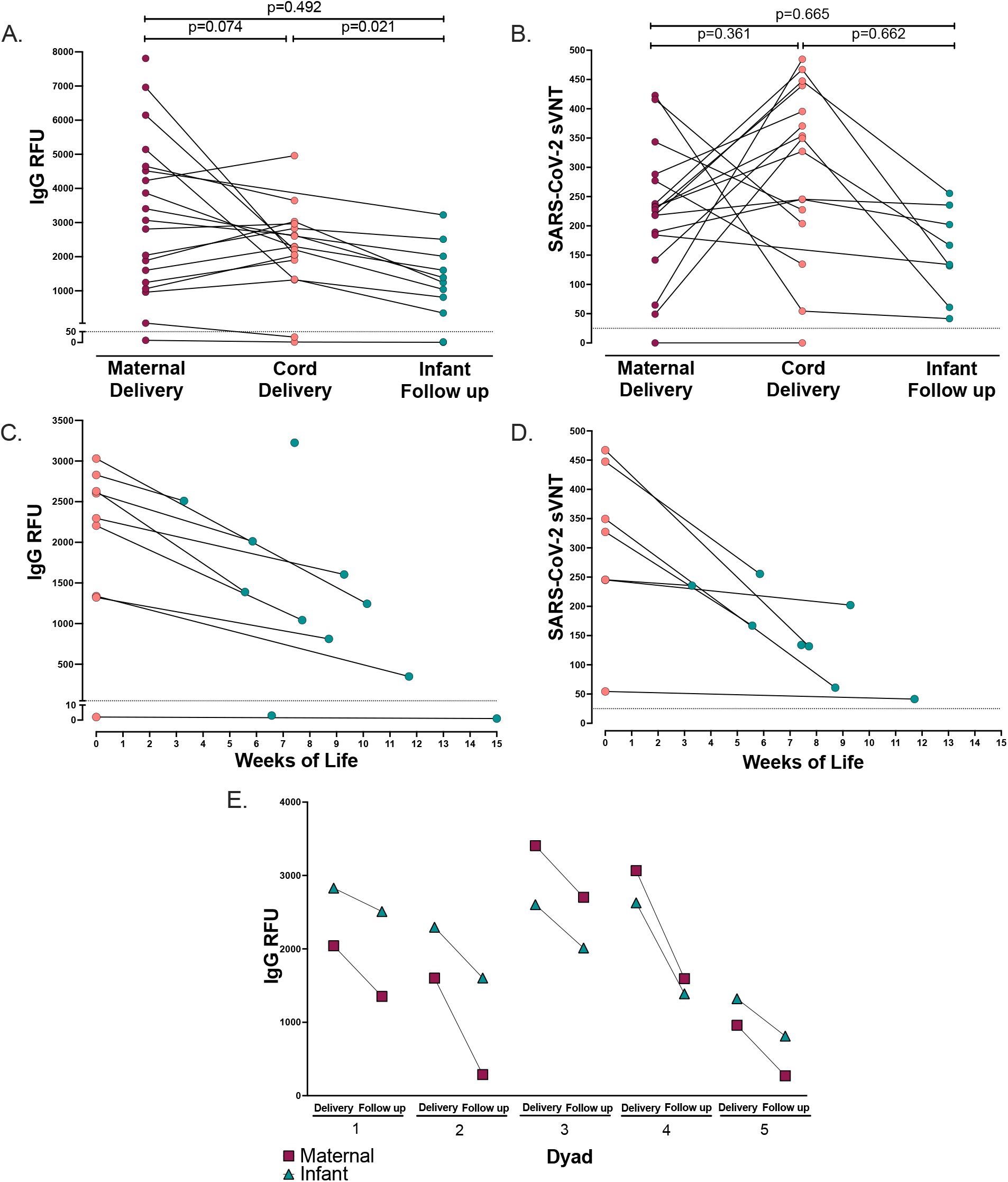
Paired maternal, cord, and infant IgG and neutralization antibodies. A. Paired maternal plasma at delivery (n=19), cord plasma (n=17), and infant follow-up (n=10) by anti-SARS-CoV-2 IgG antibody relative fluorescence units (RFU), (Spearman’s rank correlation, dotted line indicates positive cutoff value of 50 RFU). B. Paired maternal plasma at delivery (n=17), cord plasma (n=16), and infant follow-up (n=8) by SARS-CoV-2 label-free surrogate neutralization assay (sVNT), (Spearman’s rank correlation, dotted line indicates positive cutoff value of 25). C. Paired cord plasma (n=9) and infant follow-up plasma (n=11) anti-SARS-CoV-2 IgG by weeks of life. D. Paired cord plasma (n=7) and infant follow-up plasma (n=8) label-free surrogate neutralization assay (sVNT) by weeks of life. E. Paired maternal plasma at delivery (n=5), cord plasma (n=5), and paired maternal follow-up (n=5) and infant follow-up plasma (n=5) anti-SARS-CoV-2 IgG. Two-sided *p* values were calculated for all test statistics.

We next evaluated the transplacental transfer of neutralizing antibody titers by a label-free surrogate neutralization assay (sVNT) from mother to cord blood. Maternal and cord blood at delivery had robust neutralizing responses (maternal n=17, mean 220.2, range [0, 422]. Cord blood n=16, mean 296.6, range [0, 485], Fig 2B). All mother-infant dyads with positive IgG serology at delivery had detectable transplacental transfer of neutralizing antibodies with the exception of one pair in which the mother was borderline IgG positive at delivery and cord blood was negative, for which both maternal and cord blood were negative for neutralizing titers (Fig 2B). However, maternal and cord blood neutralizing titers were not significantly correlated (p=0.361, R_s_=-0.244, Fig 2B). Taken together, this indicates that maternal mRNA vaccination induces functional neutralizing antibodies that are transferred to the infant.

### Maternally-derived vaccine induced anti-SARS-CoV-2 IgG and neutralizing antibodies persist through early infancy

A subset of infants was sampled at convenience timepoints during follow up (infant n=11, weeks of life range [3,15] mean 8.3 weeks). Anti-SARS-CoV-2 IgG levels were positive in 81.8% of infants at follow-up (9/11 infants, mean 1290, range [1, 3225] RFU, Fig 2A), with one infant still IgG positive at 12 weeks of age (Fig 2C). The two infants that were IgG negative at follow up were both born to mothers who received only one vaccine dose prior to delivery (6 and 9 days, respectively). One of these infants did not have paired maternal or cord blood available at the time of delivery for comparison, and the other was IgG negative in cord blood. Maternal and infant follow-up anti-SARS-CoV-2 IgG levels were not significantly correlated; however, cord blood and infant follow-up IgG levels were significantly associated (p=0.492, R_s_=0.249 and p=0.021, R_s_=0.76, respectively, Fig 2A). All infants were IgM negative at the time of follow up.

All infants with available IgG positive samples at follow up had detectable neutralizing titers (n=8, mean 154, range [41-256], Fig 2B). Maternal and infant follow-up neutralizing titers were not significantly correlated, as well as cord and infant follow up neutralizing titers (p=0.665, R_s_=-0.191 and p=0.662, R_s_=0.214, respectively, Fig 2B).

To compare the rate of decay of IgG antibody levels in mothers and their infants, we evaluated 5 dyads with paired maternal and infant blood samples on the same day at the time of follow-up (range 3-9 weeks post-delivery). Maternal antibody IgG levels decreased faster in mothers than infants (mean delta -974 RFU and -670 RFU, respectively. Fig 2E) at the follow up timepoint. Taken together this indicates, maternally-derived functional vaccine induced antibodies persist at high levels in newborns through early infancy during a critical time of immune vulnerability and may decay slower than maternal IgG antibodies.

### Vaccine induced antibody timing and transplacental facilitated transfer

We assessed the relationship of anti-SARS-CoV-2 IgG levels to neutralizing antibody levels. We found a strong correlation between IgG and neutralizing titers in maternal plasma at delivery (R_s_=0.744, p=0.0012) and infant follow up (R_s_=0.738, p=0.046) timepoints, but no significant association between IgG and neutralizing titers in cord blood (R_s_=0.121, p=0.656, Figure 3).

**Figure 3.**
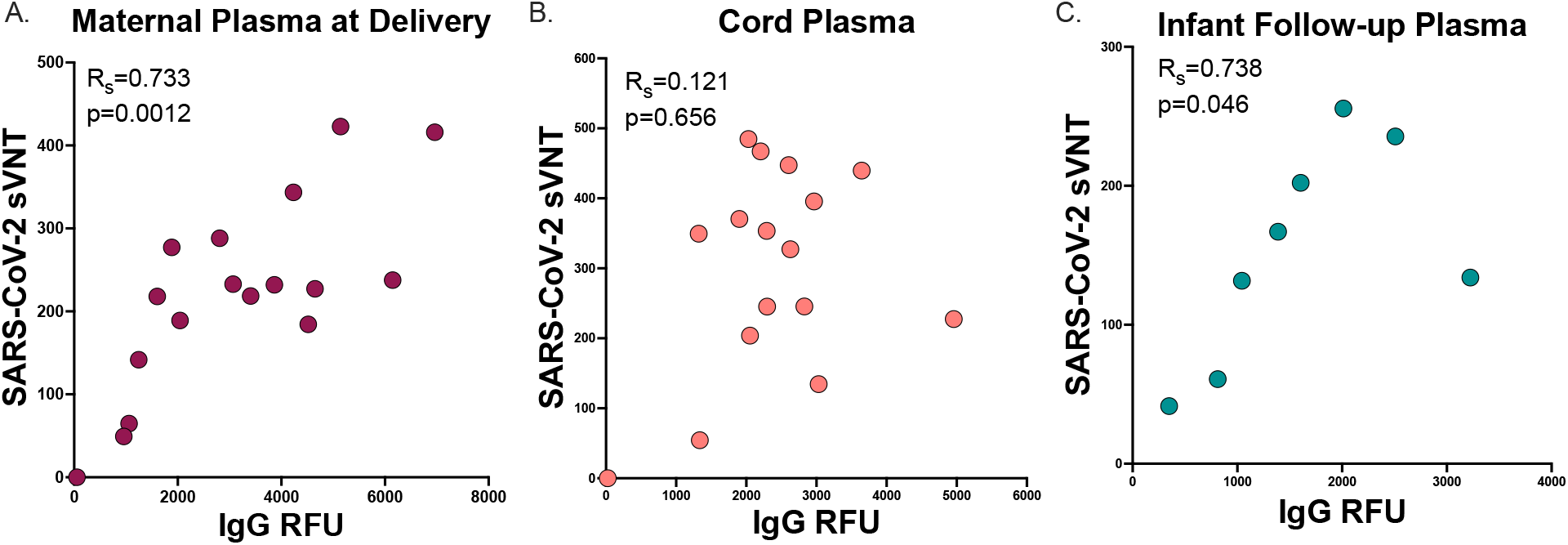
Neutralization to IgG antibody correlation. A. Maternal plasma at delivery (n=17) B. Cord plasma (n=16) C. Infant follow-up plasma (n=8) SARS-CoV-2 label-free surrogate neutralization assay (sVNT) by anti-SARS-CoV-2 IgG correlation (Spearman’s rank correlation). Two-sided *p* values were calculated for all test statistics.

We then evaluated the impact of timing of vaccination on maternal antibody levels at delivery. We found no statistically significant correlation between maternal IgG levels at delivery and time since dose 1 (R_s_=-0.335, p=0.160) and gestational age at delivery (R_s_=0.270, p=0.265, Fig 4A,B). This lack of correlation appeared to be driven by two participants that had low or absent levels of antibodies at delivery and received their first dose of vaccine within 30 days of delivery. We then evaluated neutralizing titers in those participants with known detectable IgG levels at delivery and found that maternal neutralizing titers at delivery trended with days since vaccine dose 1 but was not statistically significantly (R_s_=-0.422, p=0.093), and maternal neutralizing titers at delivery was not associated with gestational age at dose 1 (R_s_=0.074, p=0.780). One participant was borderline IgG positive at delivery (vaccinated within 30 days of delivery) and did not have detectable neutralizing titers at delivery (Fig 4C,D).

**Figure 4.**
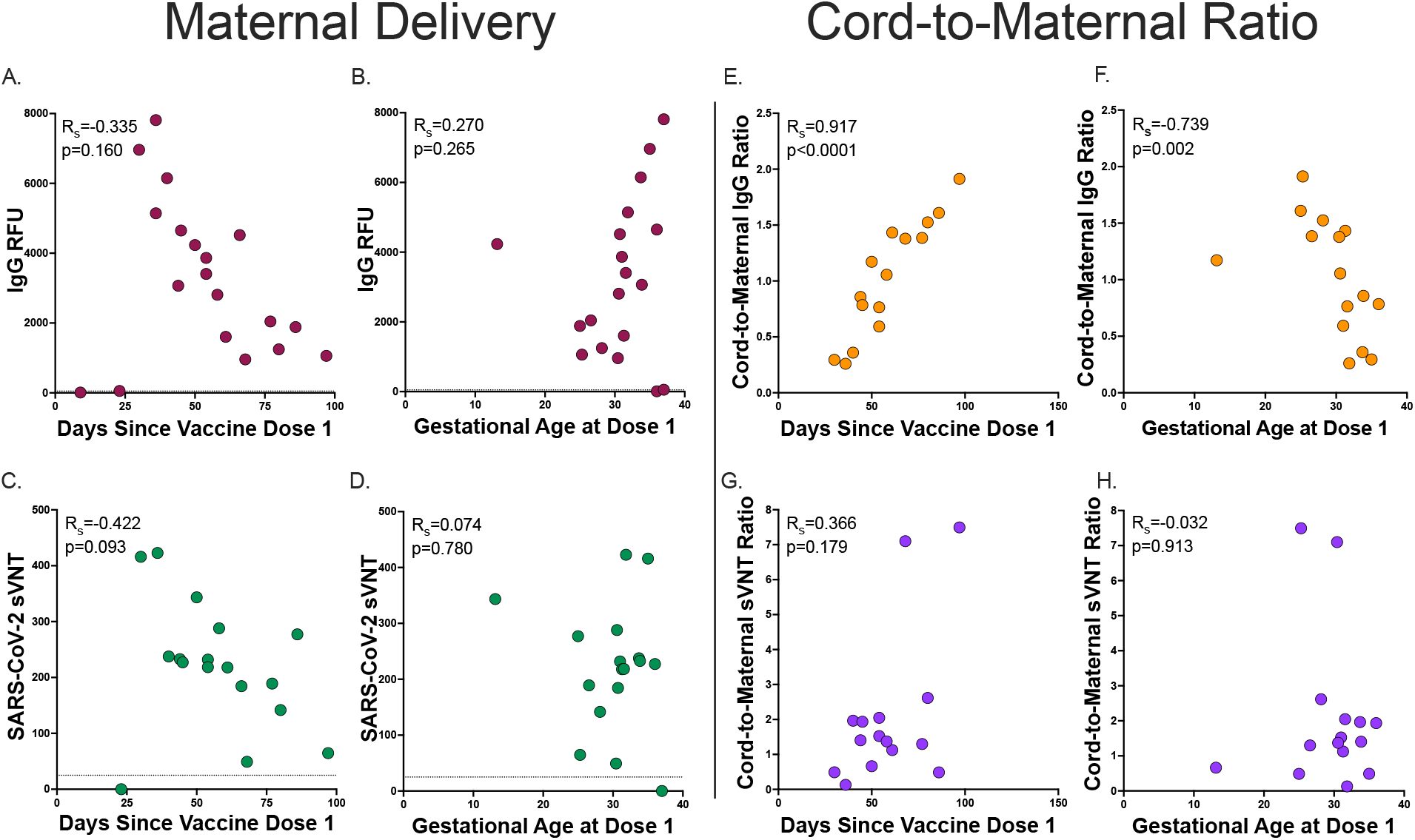
Maternal delivery and Cord-to-maternal antibody transfer ratios timing. A. Maternal delivery anti-SARS-CoV-2 IgG antibody transfer ratio by days since vaccine dose 1 (n=19, dashed line indicates positive cutoff >50 RFU) B. Maternal delivery anti-SARS-CoV-2 IgG antibody transfer ratio by gestational age at vaccine dose 1 (n=19, dashed line indicates positive cutoff >50 RFU) C. Maternal delivery SARS-CoV-2 label-free surrogate neutralization assay (sVNT) antibody transfer ratio by days since vaccine dose 1 (n=17, dashed line indicates positive cutoff >25). D. Maternal delivery SARS-CoV-2 label-free surrogate neutralization assay (sVNT) antibody transfer ratio by gestational age at vaccine dose 1 (n=17, dashed line indicates positive cutoff >25) E. Cord-to-maternal anti-SARS-CoV-2 IgG antibody transfer ratio by days since vaccine dose 1 (n=15) F. Cord-to-maternal anti-SARS-CoV-2 IgG antibody transfer ratio by gestational age at vaccine dose 1 (n=15) G. Cord-to-maternal SARS-CoV-2 label-free surrogate neutralization assay (sVNT) antibody transfer ratio by days since vaccine dose 1 (n=15). H. Cord-to-maternal SARS-CoV-2 label-free surrogate neutralization assay (sVNT) antibody transfer ratio by gestational age at vaccine dose 1 (n=15). Two-sided *p* values were calculated for all test statistics.

To assess facilitated antibody transfer, we evaluated cord-to-maternal antibody IgG and neutralization titer ratios by time since vaccination and gestational age. We found that IgG ratios were highly correlated with both time since first maternal vaccination dose and gestational age at first dose (R_s_=0.917, p<0.0001 and R_s_=-0.739, p=0.002, respectively. Fig 4E,F). However, neutralization titer cord-to-maternal ratios by time since first vaccination dose and gestational age at first dose were not significantly associated (R_s_=0.366, p=0.179 and R_s_=-0.032, p=0.913, respectively, Figure 4G,H). Together, this may indicate that timing of vaccination in pregnancy is critical for maternal-fetal antibody transfer, and functional neutralizing antibodies are differentially transferred to the fetus as compared to total anti-SARS-CoV-2 IgG during gestation.

### mRNA vaccination leads to a unique SARS-CoV2 Spike protein antibody epitope binding signature

We next investigated antibody linear epitope binding and transplacental transfer using the PhIP-seq/VirScan SARS-CoV-2 Spike protein phage display array in mother-infant dyads at the time of birth (Figure 5). We found that timing of vaccination was important for the trans-placental transfer of Spike protein epitope binding antibodies. Two mother-infant dyads had minimal Spike protein specific epitope binding. The first dyad only received one dose of mRNA vaccine 9 days prior to delivery, and the other dyad received the second vaccine dose 2 days prior to delivery.

**Figure 5.**
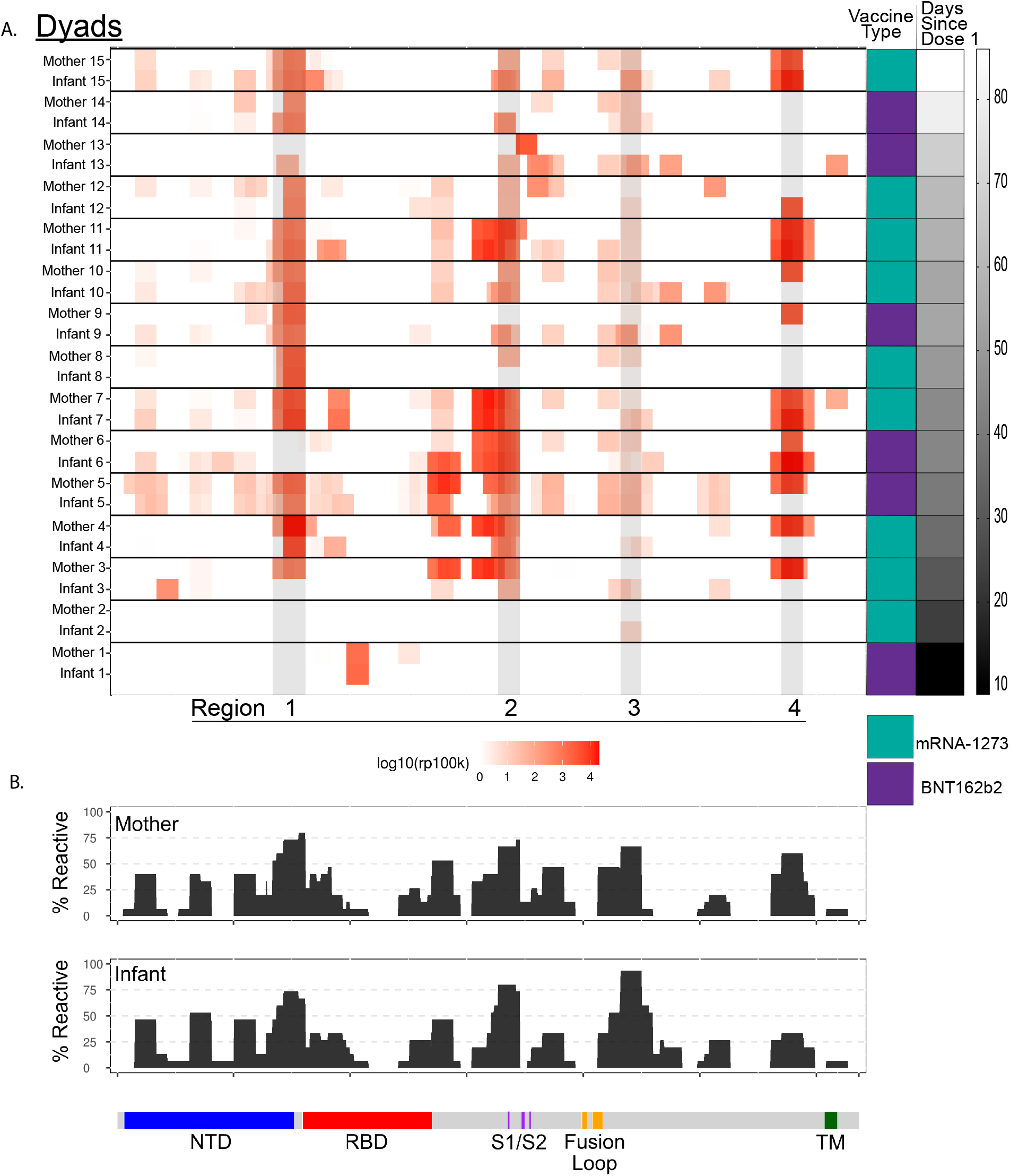
PhIP-seq/VirScan paired maternal and cord SARS-CoV-2 Spike protein epitope binding. A. Heatmap displaying results of significant enriched (p<0.001) linear SARS-CoV-2 Spike protein epitope binding from 15 paired mother-infant dyads in maternal plasma at delivery and cord plasma by vaccine type and time since vaccine dose 1. Areas of high cumulative epitope binding designated by regions 1-4. B. Cumulative fold enrichment of mothers and infants linear SARS-CoV-2 Spike protein epitope binding.

We found high levels of SARS-CoV-2 Spike protein epitope binding in 4 major peaks we designate as regions 1-4 (Figure 5A). Region 1 overlays the carboxy terminal of the N-terminal domain. Region 2 overlaps with key residues for the S1/S2 cleavage site. Regions 3 and 4 are within S2, flanking the fusion loop and the transmembrane portion of the Spike protein, respectively. However, we found minimal binding in the receptor binding domain (RBD) of Spike protein. Prior evaluation using the PhIP-seq/VirScan SARS-CoV-2 epitope phage array during SARS-CoV-2 infection demonstrated similar binding in regions 3 and 4, however in SARS-CoV-2 infection there was minimal binding in regions 1 and 2 demonstrating that antibody epitope binding in these regions may be unique to vaccination^18^. Additionally, there is proportional representation of linear epitope binding across the SARS-CoV-2 Spike protein proteome between mothers and infants (Figure 5B). Taken together, SARS-CoV-2 antibody linear epitope binding after vaccination during pregnancy shows similar patterns, with multiple immunodominant regions found in the majority of mothers and infants. Some of these regions are unique to vaccination and not observed during natural infection^18-20^.

## Discussion

Among twenty women who received the COVID-19 mRNA vaccine during pregnancy, our study found no evidence of transplacental transfer of mRNA vaccine products but did find high levels of functional vaccine-derived antibodies that transferred to the infant at delivery and persisted during early infancy. Additionally, we identified high levels of epitope binding in two regions of Spike protein unique to SARS-CoV-2 vaccination^18^. These data may address some of the many unanswered questions regarding COVID-19 vaccination in pregnancy: including the dynamics of antibody production in the pregnant immune state, and the optimal timing of immunization in pregnancy to impart passive immunity to the newborn during the vulnerable first few weeks of infancy.

Uptake of COVID-19 vaccination in pregnancy has been slow^5^, and reasons for vaccine hesitancy are likely multifactorial — but theoretical concerns that vaccine mRNA could cross the placenta have been raised. We found no evidence of mRNA vaccine products in any of our delivery samples. Additionally, no infants in our study had a fetal immune response to Spike protein as demonstrated by a negative anti-SARS-CoV-2 IgM antibody in cord blood and infant follow up samples. This further supports the lack of transfer of vaccine products, as only IgG is transferred from the mother, and IgM production would indicate an endogenous fetal immune response which has rarely been seen in natural infection with SARS-CoV-2 during pregnancy ^16,21-23^. This provides additional reassurance that mRNA vaccination is safe during pregnancy.

We found that the timing of immunization during pregnancy is important to ensure trans-placental transfer of protective antibodies to the neonate, and during critical windows of immune vulnerability during early infancy. Consistent with prior studies showing robust immune responses to mRNA vaccination^14,15,17^, we found high levels of IgG after two doses of mRNA vaccine. However, completion of the vaccination series well before delivery was important to ensure transfer of antibodies to the infant. Two mothers only received one vaccine dose prior to delivery and did not transfer antibodies as demonstrated by the lack of antibodies in cord (in one with available cord blood) and in both infants at follow-up. Additionally, neutralizing antibodies were not transferred in a mother who received her second dose of vaccine 2 days prior to delivery. All evaluated mothers who received both doses during pregnancy and with the second dose greater than 9 days prior to delivery transferred IgG and neutralizing antibodies to their infants. Consistent with early studies of antibody transfer after COVID-19 vaccination in pregnancy, most of our participants were vaccinated in the third trimester of pregnancy. Larger studies of individuals vaccinated prior to pregnancy and in the first and second trimester are needed to understand persistence and waning of vaccine-induced immune responses.

Additionally, we believe we are the first to report that infants in the first few months of life continued to have maternal vaccine-derived anti-SARS-CoV-2 antibodies that were functional as demonstrated by high levels of neutralizing antibodies presenting infants up to 12 weeks of age. This is consistent with known persistence of maternally-derived antibodies from other vaccinations including pertussis, rubella, varicella^24-26^. Additionally, we have previously found persistence of anti-SARS-CoV-2 IgG antibodies in infants after natural infection up to 6 months^16^. However, the functional capability of these antibodies as compared to anti-SARS-CoV-2 vaccination-derived antibodies is unknown. Further evaluation of the longitudinal persistence of maternal vaccine-derived antibodies during infancy will be critical to determine optimal timing of COVID-19 vaccination in infancy.

Evaluation of paired maternal and baby samples at post-partum follow up timepoints showed a faster decline in maternal IgG antibody levels than infants, suggesting that persistence of maternally-derived antibody may be prolonged for infants. Differences in renal excretion and neonatal Fc receptor (FcRn) expression, which is involved in antibody degradation^27^ in the infant as compared to adults, could underly these differences and should be explored further.

Consistent with observations in non-pregnant adults, we found that IgG levels in mothers at delivery, and at infant follow-up were highly correlated with neutralizing titers^28^. However, cord blood IgG levels did not correlate with neutralizing titers. Moreover, IgG cord-to-maternal ratios, which represent a proxy of maternal to fetal antibody transfer, were highly correlated with timing of vaccination (gestational age and days since the first dose), but cord-to-maternal neutralizing titer ratios were not significantly associated with time since vaccination nor gestational age. During gestation there is facilitated transfer of maternally derived antibodies through the binding of the neonatal Fc receptor in the synctiotrophoblast layer^29^. Differences in glycosylation^30,31^, FcR/FcRn binding affinity^17,32^, preferential IgG subclass transfer^33,34^ may be different in functional neutralizing antibodies as compared to total IgG antibody transfer.

However, a limitation of this study is the majority of participants were vaccinated in the third trimester. Further investigations on factors that influence the transport of functional antibodies across trimesters are needed to understand antibody dynamics and optimal transfer of protective antibodies to infants.

Using a PhIP-seq/VirScan SARS-CoV-2 Spike protein phage array we were able to compare linear epitope antibody binding in mothers and their infants. Consistent with IgG and neutralizing antibody evaluation, timing of vaccination was critical to ensure the transplacental transfer of antibodies to the infant. Additionally, we identified unique regions of antibody epitope binding in our vaccinated cohort that were not identified using the same phage library in a prior evaluation of a cohort of SARS-CoV-2 infected individuals^18^. One of these regions included the carboxy terminal of the N-terminal domain, with other work having shown that the N-terminal domain is targeted by neutralizing antibodies against Spike protein^35^. We did not see significant binding in the receptor binding domain (RBD), which may be attributable to the fact that the phage display library displayed short, linear peptides while antibodies targeting RBD are known to target conformational epitopes. Lastly, we found that the same immunodominant regions targeted by antibodies targeting the Spike protein in both mothers and infants.

In summary, this work provides further evidence that mRNA vaccination is safe in pregnancy and demonstrates that it generates time-dependent protective, functional antibody responses in mothers and infants that persist during early infancy.

## Methods

### Cohort and Sample collection

The University of California San Francisco (UCSF) institutional review board approved the study (20-32077). Written informed consent was obtained from all participants. We enrolled 20 pregnant individuals who were vaccinated with either BNT-162b2 or mRNA-1273 mRNA vaccines. Pregnant individuals were followed through delivery, and their infants were followed up to 12 weeks of life. Maternal blood was collected during pregnancy (pre-vaccine, 3-4 weeks post-dose 1, 4-8 weeks post-dose 2). During delivery, maternal blood, placenta tissue, and cord blood was collected. Infant follow-up blood was collected at convenience timepoints. Whole blood was immediately added to RNAlater in a 1:1.3 ratio. Plasma was isolated from whole blood by centrifugation and immediately cryopreserved. Full-thickness placental biopsy was collected within 1 hour of delivery, washed three times with phosphate buffered saline, and preserved in RNAlater.

### SARS-CoV-2 plasma serology

Anti-SARS-CoV-2 plasma IgM and IgG antibodies were measured using the Pylon 3D automated immunoassay system (ET Healthcare, Palo Alto, CA). In brief, quartz glass probes are pre-coated with either affinity purified goat anti Human IgM (IgM capture) or Protein G (IgG capture) are dipped into diluted patient sample. Samples are washed, and then the probe is dipped into the assay reagent containing both biotinylated recombinant spike protein receptor binding domain (RBD) and nucleocapsid protein (NP). After a washing, the probe is incubated with a Cy®5-streptavidin (Cy5-SA) polysaccharide conjugate reagent, allowing for cyclic amplification of the fluorescence signal. The background corrected signal is reported as relative fluorescent units (RFU) which is proportional to the amount of specific antibodies in the sample allowing for quantification. Levels of IgM and IgG were considered positive if greater than 50 relative fluorescence units.

### SARS-CoV-2 neutralizing assay

SARS-CoV-2 antibody neutralization titers were measured using a label-free surrogate neutralization assay (LF-sVNT) previously described^28^. Briefly, the method measures the binding ability of recombinant RBD (Sino Biological, Wayne, PA) coated onto sensing probes (Gator Bio, Palo Alto, CA) to recombinant ACE2 (Sino Biological, Wayne, PA) after neutralizing RBD with SARS-CoV-2 antibodies in serum. Measurements were done using a thin-film interferometry (TFI) label-free immunoassay analyzer (Gator Bio, Palo Alto, CA). Each serum sample was diluted in a series (1:25, 1:100, 1:250, 1:500, 1:1000, 1:2000) in running buffer (PBS at pH 7.4 with 0.02% Tween 20, 0.2% BSA, and 0.05% NaN_3_) for analysis. The first testing cycle for each diluted sample measured the binding ability of RBD to ACE2 with neutralization, and the second cycle provided the full binding ability of RBD without neutralization. In each cycle, the recorded time course of signals, as known as the sensorgram, was recorded. The readout measured the signal increase in RBD-ACE2 complex formation, representing the quantity of RBD-ACE2 complex on the sensing probe. A neutralization rate was calculated as the ratio of the readout in the first cycle to that in the second cycle, presented as a percentage. To obtain the neutralizing antibody titer (IC_50_) for each serum sample, the neutralization rates were plotted against dilutions, and the points were fitted using a linear interpolation model. The reciprocal of the dilution resulting in a 50% neutralization rate was defined as the neutralizing antibody titer.

### SARS-CoV-2 Spike protein Western blot

Maternal blood and cord blood were diluted in RNAlater in 1:1.3 ratio, placenta was preserved in RNAlater. Protein lysates were obtained from samples using RIPA buffer (150 mM NaCl, 25 mM Tris-HCl (pH 7.4), 1% NP-40, 0.5% sodium deoxycholate, 0.1% sodium dodecyl sulfate) containing Halt™ protease inhibitor cocktail (ThermoScientific).Cell Lysates were resolved by SDS/PAGE on a Bis-Tris methane 4–12% polyacrylamide gel and transferred to a nitrocellulose membrane, blocked with 5% skimmed milk diluted in PBS, an incubated overnight at 4 C with anti-SARS-CoV-2 Spike mouse mAb (1A9, GeneTex) or anti-GAPDH rabbit polyclonal antibody (GT×100118, GeneTex) respectively diluted 1:1,000 or 1:5,000 in blocking buffer. The membrane was washed in PBS buffer containing Tween-20 (0.1%) and then incubated for 1 h with horseradish peroxidase-conjugated anti-mouse and anti-rabbit secondary antibody (Jackson ImmunoResearch) diluted respectively 1:5,000 and 1:10,000. The membrane was thoroughly washed, and proteins visualized using Immobilon Forte Western HRP substrate (Millipore).

### SARS-CoV-2 Spike mRNA PCR

Maternal blood and cord blood were diluted in RNAlater in 1:1.3 ratio, placenta was preserved in RNAlater. Tissues were kept at -80°C until analyzed. RNA was isolated from samples using the RNeasy Micro or Mini Kit (Qiagen) according to manufacturer’s protocol. RNA concentration was measured using nanodrop and all samples had >30 ng/ul total RNA. 500ng RNA was transcribed into cDNA using qScript cDNA synthesis kit (Quantabio). Primers were design to detect the vaccines mRNA (mRNA-1273 Moderna and BNT162b2 Pfizer-BioNtech) as previously described^36^. Forward primer: AACGCCACCAACGTGGTCATC. Reverse primer: GTTGTTGGCGCTGCTGTACAC. Primers were shown to detect samples containing as low as 1.5 pg of vaccine using vaccine standard curve (Table S2). QuantaStudio 6 Flex (Applied Biosystems) instrument and SsoFast EvaGreen supermix (Bio-Rad) were used for PCR reaction: 30 second 95°C followed by 40 cycles of 5 second 95°C and 20 seconds 60°C. All samples were run in triplicate as 20 μL reactions, and Ct values corresponding to <1.5 pg of vaccine based on standard curve (Table S2) were interpreted as a negative result. For vaccines cDNA standard curves, 10000 pg/μL vaccine mRNA (as cDNA) sample was used for serial dilution in 1:3 ratio, up to 0.06 pg/μL. Two μL of these diluted samples were used in each well to create standard curves.

### PhIP-Seq/VirScan Coronavirus phage display assay

#### Immunoprecipitation of phage-bound patient antibodies

Maternal plasma at delivery and cord plasma were evaluated by PhIP-Seq/Virscan Coronavirus phage display. Construction of the Coronavirus PhIP-Seq library and detailed methods for immunoprecipitation, sequencing and bioinformatic processing of data are identical to what has previously been described^18^. For the purposes of the analysis conducted in this study, analysis was restricted to sero-reactivity against the SARS-CoV-2 Spike protein. As previously described, a total of two rounds of amplification and selection were performed for all PhIP-Seq analyses.

#### Next Generation Sequencing library prep

Amplicon sequencing library preps were performed using the Labcyte Echo 525 and an Integra Via Flow 96 and were identical to what has previously been described^18^. All libraries were pooled by equal volume, cleaned and size selected using Ampure XP beads at 1.0X per manufacturer’s protocol. Libraries were quantified by High Sensitivity DNA Qubit and quality-checked by High Sensitivity DNA Bioanalyzer. Sequencing was then performed on a NovaSeq S1 (300 cycle kit with 1.3 billion clusters) aiming for sequencing depths of at least 1 million reads per sample.

#### Bioinformatic Analysis of PhIP-Seq Data

Sequencing reads were aligned to a reference database of the full viral peptide library using the Bowtie2 aligner. For all VirScan libraries, the null distribution of each peptide’s log10(rpK) was modeled using a set of 95 pre-pandemic, healthy control sera. All counts were augmented by 1 to avoid zero counts in the healthy control sera samples. Multiple distribution fits were examined for these data, with the Normal distribution showing the best fit. These null distributions were used to calculate p values for the observed log10(rpK) of each peptide within a given sample. The calculated p values were corrected for multiple hypothesis using the Benjamini-Hochberg method. Any peptide with a corrected p value of < 0.001 was considered significantly enriched over the healthy background. To identify regions targeted by host antibodies, all library peptides were aligned to the SARS-CoV-2 reference genome. Using the aligned position of the significantly enriched peptides which aligned full-length against the reference, we determined the proportion of individuals (mothers and infants) that were reactive at each residue of the Spike protein. All plots were generated using the R ggplot2 package.

### Statistical analysis

Statistical analyses were performed using PRISM v9.2 (GraphPad), STATA 16 (StataCorp), and R version 3.6.3 and R Studio version 1.1.447. Descriptive statistics include mean, standard deviations, and ranges for continuous variables. The Wilcoxon rank-sum test was used for two-group comparisons of continuous variables including maternal pre-and post-vaccine antibody responses. Associations between continuous variables were assessed using Spearman’s rank correlation (R_s_) including comparisons between maternal, cord and infant follow-up antibody IgG and neutralizing titer responses, and timing of vaccination. Two-sided *p* values were calculated for all test statistics, and *p*<0.05 was considered significant. PhIP-Seq/VirScan bioinformatics as detailed above.

## Data Availability

All data produced in the present study are available upon reasonable request to the authors

## Data Availability

The data set generated during and/or analyzed during the current study are available from the corresponding author on reasonable request.

## Acknowledgements

M.P. was supported by the National Institutes of Health (NIAID K23AI127886), the Marino Family Foundation, and UCSF REAC award. Y.G. was supported by the Weizmann Institute of Science -National Postdoctoral Award Program for Advancing Women in Science, and of the International Society for Research in Human Milk and Lactation (ISRHML) Trainee Bridge Fund. Y.M. and W.C.G. were supported by The Roddenberry Foundation. S.L.G. was supported by the National Institutes of Health (NIAID K08AI141728), and the Bill and Melinda Gates Foundation (INV-017035), and a generous gift from the Kryzewski Family.

We thank all the mothers and infants that participated in this study. We thank Kenneth Scott, BS, RPh, (UCSF Health Pharmacy, University of California, San Francisco) and Hannah J. Jang, PhD, RN, PHN, CNL (UCSF School of Nursing, University of California, San Francisco), for voluntarily providing unused vaccine for this study, and to Dr. Margaret Feeney (University of California, San Francisco) and Dr. Nadav Ahituv (University of California, San Francisco) for support of these experiments.

## Author contributions

M.P. Helped conceive and design the project, oversaw recruitment, designed, and performed sample collection, oversaw experiment design, oversaw data analysis, provided funding, and drafted the manuscript.

Y.G. Recruited and consented enrollees, oversaw sample collection, designed, performed, and analyzed mRNA PCR experiments, performed data analysis.

A.G.C. Recruited and consented enrollees, oversaw sample collection, performed chart review, and helped draft the manuscript.

Y.M. Performed and helped design Western blot.

L.L. Performed and analyzed mRNA PCR experiments, performed sample collection.

B.A. Performed phage immunoprecipitation assays.

H.C. and U.J. performed and helped design critical experiments, and data collection.

C.Y.L., V.J.L., M.C., L.W., S.B. Performed and coordinated sample collection, and data collection.

V.J.F. Helped conceive and coordinate the project.

A.P.M. Provide funding.

W.C.G. Helped design western blot and oversaw data analysis.

A.H.B.W Designed and oversaw all serology experiments.

K.L.L. Designed and oversaw all neutralizing antibody experiments.

J.R. Designed, analyzed, and oversaw phage immunoprecipitation sequencing assays.

S.L.G. conceived and designed the project, oversaw recruitment, oversaw experiment design, oversaw data analysis, provided funding, and helped draft the manuscript.

M.P., Y.G., Y.U., L.L., A.H.B.W, W.C.G, K.L.L., and S.L.G verified the underlying data.

All authors reviewed and approved the manuscript.

