## Supplemental Materials for "Evaluation of transplacental transfer of mRNA vaccine products and functional antibodies during pregnancy and early infancy"

Supplementary Material

Table of Contents

| | Page |
|---|---|
| Table S1 | 2 |
| Table S2 | 2 |
| Table S3 | 3 |
| Figure S1 | 4 |

**Table S1: Maternal Demographics**

| Study ID | Gestational Age at Delivery | Infant Sex | Vaccine Type | Gestational Age at Dose 1 |
|---|---|---|---|---|
| 11012 | 37.71 | Female | Moderna | 26.572 |
| 11018 | 37.43 | Female | Moderna | 36 |
| 11026 | 37.57 | Female | Moderna | 31.858 |
| 11033 | 20.43 | Male | Pfizer | 13.143 |
| 11036 | 38.71 | Male | Moderna | 31 |
| 11038 | 39.86 | Female | Moderna | 31.286 |
| 11040 | 39.29 | Male | Moderna | 33.715 |
| 11045 | 38.86 | Female | Pfizer | 30.572 |
| 11047 | 39.29 | Male | Moderna | 31.572 |
| 11048 | 39.14 | Male | Pfizer | 25.286 |
| 11058 | 40.14 | Female | Moderna | 33.858 |
| 11059 | 40.14 | Male | Moderna | 30.429 |
| 11062 | 40.29 | Male | Moderna | 30.715 |
| 11063 | 37.57 | Female | Moderna | 25 |
| 11085 | 41.14 | Male | Pfizer | 40.286 |
| 11091 | 39.57 | Female | Pfizer | 28.143 |
| 11123 | 39.14 | Male | Pfizer | 35 |
| 11126 | 39.14 | Female | Pfizer | 36 |
| 11128 | 39.71 | Male | Pfizer | 37 |
| 11137 | 40.57 | Male | Moderna | 37 |

**Table S2: Serial dilution of vaccine cDNA**

| pg Vaccine cDNA | Moderna Ct | Pfizer Ct |
|---|---|---|
| 10000.00 | 6.578 | 6.696 |
| 3333.33 | 8.247 | 12.266 |
| 1111.11 | 11.403 | 12.804 |
| 370.37 | 12.799 | 15.355 |
| 123.46 | 14.327 | 16.314 |
| 41.15 | 16.217 | 19.397 |
| 13.72 | 18.430 | 19.727 |
| 4.57 | 20.503 | 21.579 |
| 1.52 | 23.315 | 24.646 |
| 0.51 | 23.337 | 23.998 |
| 0.17 | 24.259 | 26.110 |
| 0.06 | 25.510 | 27.840 |

*Sensitivity to 1.5 pg/uL

**Table S3: Spike protein western blot and vaccine mRNA PCR results**

| | Spike Protein Detection (Western blot) | | | Spike mRNA Detection (qRT-PCR) | | |
|---|---|---|---|---|---|---|
| Participant | Placenta | Maternal Blood | Cord Blood | Placenta | Maternal Blood | Cord Blood |
| 11012 | Negative | Negative | Negative | Negative | Negative | Negative |
| 11018 | Negative | Negative | Negative | Negative | Negative | Negative |
| 11026 | Negative | Negative | Negative | Negative | Negative | Negative |
| 11033 | Negative | Negative | Negative | Negative | Negative | Negative |
| 11036 | Negative | Negative | Negative | Negative | Negative | Negative |
| 11038 | Negative | Negative | Negative | Negative | Negative | Negative |
| 11040 | - | Negative | Negative | - | Negative | Negative |
| 11045 | Negative | Negative | Negative | Negative | Negative | Negative |
| 11047 | - | Negative | Negative | - | Negative | Negative |
| 11048 | Negative | Negative | Negative | Negative | Negative | Negative |
| 11058 | Negative | Negative | Negative | Negative | Negative | Negative |
| 11059 | - | Negative | Negative | - | Negative | Negative |
| 11062 | Negative | Negative | - | Negative | Negative | - |
| 11063 | Negative | Negative | Negative | Negative | Negative | Negative |
| 11085 | - | - | - | - | - | - |
| 11091 | Negative | Negative | Negative | Negative | Negative | Negative |
| 11123 | Negative | Negative | Negative | Negative | Negative | Negative |
| 11126 | Negative | Negative | Negative | Negative | Negative | Negative |
| 11128 | Negative | Negative | Negative | Negative | Negative | Negative |
| 11137 | Negative | Negative | - | Negative | Negative | Negative |

- Missing sample

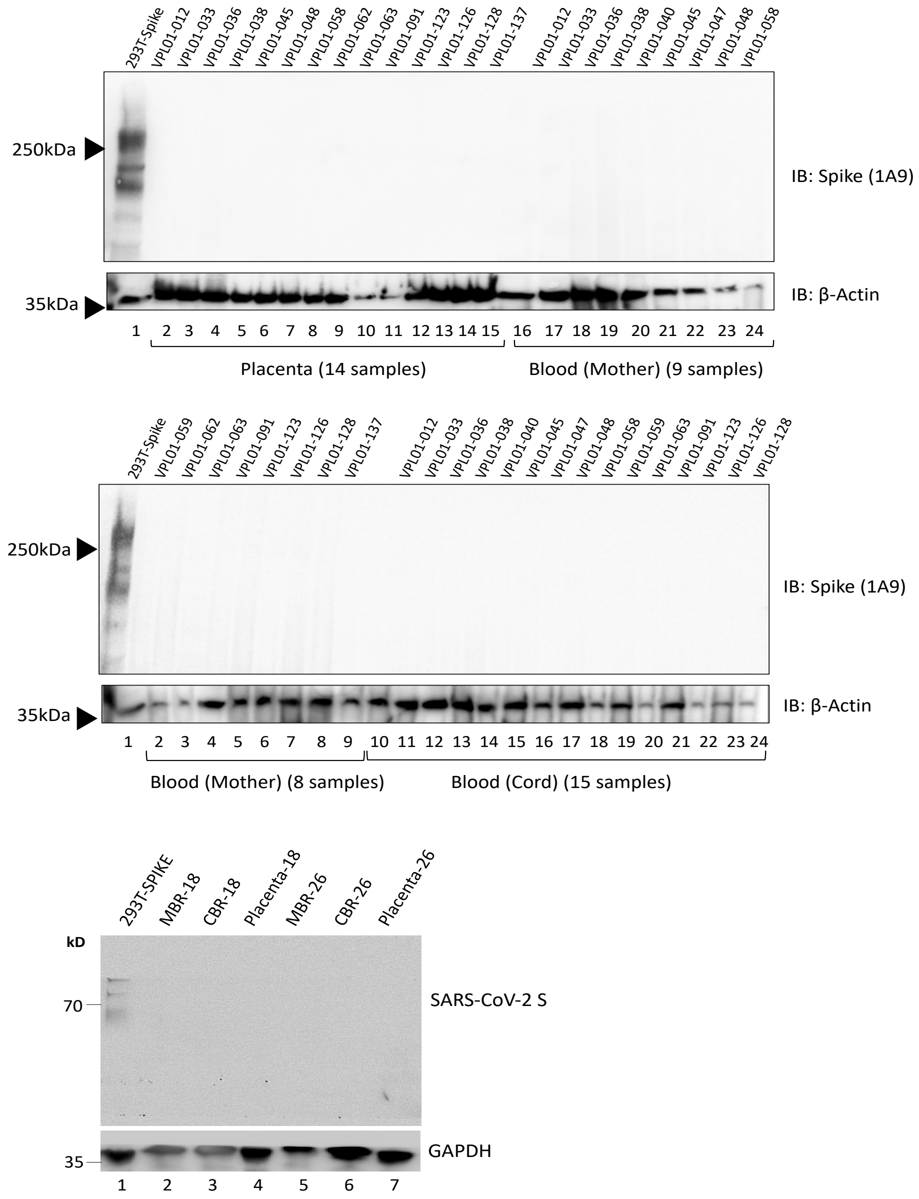


**Supplementary Figure 1: Spike protein Western blot**

Western blot of maternal blood at delivery, placenta tissue, and cord blood compared to Spike protein positive control
